# Assessment of the risk of SARS-CoV-2 reinfection in an intense re-exposure setting

**DOI:** 10.1101/2020.08.24.20179457

**Authors:** Laith J. Abu Raddad, Hiam Chemaitelly, Joel A. Malek, Ayeda A. Ahmed, Yasmin A. Mohamoud, Shameem Younuskunju, Houssein H. Ayoub, Zaina Al Kanaani, Abdullatif Al Khal, Einas Al Kuwari, Adeel A. Butt, Peter Coyle, Andrew Jeremijenko, Anvar Hassan Kaleeckal, Ali Nizar Latif, Riyazuddin Mohammad Shaik, Hanan F. Abdul Rahim, Hadi M. Yassine, Mohamed G. Al Kuwari, Hamad Eid Al Romaihi, Sheikh Mohammad Al Thani, Roberto Bertollini

**Affiliations:** Infectious Disease Epidemiology Group, Weill Cornell Medicine-Qatar, Cornell University, Doha, Qatar; World Health Organization Collaborating Centre for Disease Epidemiology Analytics on HIV/AIDS, Sexually Transmitted Infections, and Viral Hepatitis, Weill Cornell Medicine–Qatar, Cornell University, Qatar Foundation – Education City, Doha, Qatar; Department of Population Health Sciences, Weill Cornell Medicine, Cornell University, New York, New York, USA; Genomics Laboratory, Weill Cornell Medicine-Qatar, Cornell University, Doha, Qatar; Department of Genetic Medicine, Weill Cornell Medicine-Qatar, Cornell University, Doha, Qatar; Department of Mathematics, Statistics, and Physics, Qatar University, Doha, Qatar; Hamad Medical Corporation, Doha, Qatar; College of Health Sciences, QU Health, Qatar University, Doha, Qatar; Biomedical Research Center, Qatar University, Doha, Qatar; Department of Biomedical Science, College of Health Sciences, Member of QU Health, Qatar University, Doha, Qatar; Primary Health Care Corporation, Doha, Qatar; Ministry of Public Health, Doha, Qatar

**Keywords:** SARS-CoV-2, epidemiology, COVID-19, incidence, infection, reinfection, immunity, genetics

## Abstract

**Background:** Reinfection with severe acute respiratory syndrome coronavirus 2 (SARS-CoV-2) is debated. We assessed risk and incidence rate of documented SARS-CoV-2 reinfection in a large cohort of laboratory-confirmed cases in Qatar.

**Methods:** All SARS-CoV-2 laboratory-confirmed cases with at least one PCR positive swab that is ≥45 days after a first-positive swab were individually investigated for evidence of reinfection, and classified as showing *strong*, *good*, *some*, or *weak/no* evidence for reinfection. Viral genome sequencing of the paired first-positive and reinfection viral specimens was conducted to confirm reinfection. Risk and incidence rate of reinfection were estimated.

**Results:** Out of 133,266 laboratory-confirmed SARS-CoV-2 cases, 243 persons (0.18%) had at least one subsequent positive swab ≥45 days after the first-positive swab. Of these, 54 cases (22.2%) had strong or good evidence for reinfection. Median time between first and reinfection swab was 64.5 days (range: 45-129). Twenty-three of the 54 cases (42.6%) were diagnosed at a health facility suggesting presence of symptoms, while 31 (57.4%) were identified incidentally through random testing campaigns/surveys or contact tracing. Only one person was hospitalized at time of reinfection, but still with mild infection. No deaths were recorded. Viral genome sequencing confirmed four out of 12 cases with available genetic evidence. Risk of reinfection was estimated at 0.01% (95% CI: 0.01-0.02%) and incidence rate of reinfection was estimated at 0.36 (95% CI: 0.28-0.47) per 10,000 person-weeks.

**Conclusions:** SARS-CoV-2 reinfection can occur but is a rare phenomenon suggestive of a strong protective immunity against reinfection that lasts for at least a few months post primary infection.

## INTRODUCTION

The severe acute respiratory syndrome coronavirus 2 (SARS-CoV-2) has been spreading around the globe causing severe disruptions to social and economic activities [1-3]. Qatar, a peninsula in the Arabian Gulf region with a diverse population of 2.8 million [4, 5], has experienced a large epidemic with one of the highest laboratory-confirmed rates of infection at >50,000 infections per million population [6, 7]. Antibody testing and mathematical modeling indicated that about half of the population of Qatar has already been infected [6].

The intensity of the epidemic with a high risk of re-exposure to the infection, as well as the availability of a centralized data-capture system of all laboratory-confirmed infections, provided an opportunity to epidemiologically assess the presence and incidence of reinfections; a debated feature of SARS-CoV-2 epidemiology whose elucidation is critical to inform global response, timing and intensity of future cycles, and impact and durability of potential vaccines [8-11].

Our aim was to assess the risk and incidence rate of documented reinfection in a cohort of 133,266 SARS-CoV-2 laboratory-confirmed infected persons. Since the relevant underlying question is whether risk of reinfection is appreciable or not, we implemented a conservative epidemiological approach for assessing documented reinfections, that is prone to overestimate rather than underestimate risk of reinfection. However, we also conducted sensitivity analyses implementing more stringent criteria for assessing reinfection. We further performed viral genome sequencing to confirm the reinfections.

## METHODS

### Sources of data

We analyzed the centralized and standardized national SARS-CoV-2 testing and hospitalization database compiled at Hamad Medical Corporation (HMC), the main public healthcare provider and the nationally-designated provider for Coronavirus Disease 2019 (COVID-19) healthcare needs. The database covers all SARS-CoV-2 cases in Qatar and encompasses data on all polymerase chain reaction (PCR) testing conducted from February 28-August 12, 2020, including testing of suspected SARS-CoV-2 cases and traced contacts and infection surveillance testing. The database further includes data on hospital admission of COVID-19 patients and the World Health Organization (WHO) severity classification for each infection [12], which is assessed through individual chart reviews by trained medical personnel. Recently, data on serological testing for antibody on residual blood specimens collected for routine clinical care from attendees at HMC were also incorporated [6].

### Laboratory methods

All PCR testing was conducted at HMC Central Laboratory or at Sidra Medicine Laboratory, following standardized protocols. Nasopharyngeal and/or oropharyngeal swabs (Huachenyang Technology, China) were collected and placed in Universal Transport Medium (UTM). Aliquots of UTM were: extracted on the QIAsymphony platform (QIAGEN, USA) and tested with real-time reverse-transcription PCR (RT-qPCR) using the TaqPath™ COVID-19 Combo Kit (Thermo Fisher Scientific, USA) on a ABI 7500 FAST (ThermoFisher, USA); extracted using a custom protocol [13] on a Hamilton Microlab STAR (Hamilton, USA) and tested using the AccuPower SARS-CoV-2 Real-Time RT-PCR Kit (Bioneer, Korea) on a ABI 7500 FAST; or loaded directly to a Roche cobas® 6800 system and assayed with the cobas® SARS-CoV-2 Test (Roche, Switzerland). The first assay targets the S, N, and ORF1ab regions of the virus; the second targets the virus’ RdRp and E-gene regions; and the third targets the ORF1ab and E-gene regions.

Serological testing was performed using the Roche Elecsys^®^ Anti-SARS-CoV-2 (Roche, Switzerland), an electrochemiluminescence immunoassay that uses a recombinant protein representing the nucleocapsid (N) antigen for the determination of antibodies against SARS-CoV-2. Qualitative anti-SARS-CoV-2 results were generated following the manufacturer’s instructions (reactive: cutoff index ≥1.0 vs. non-reactive: cutoff index <1.0).

### Inclusion criteria

All SARS-CoV-2 laboratory-confirmed cases with at least one PCR positive swab that is ≥45 days after a first-positive swab were considered as *suspected cases* of reinfection. The 45-day cutoff was informed by data from observational cohorts of SARS-CoV-2 infected persons [14, 15], and was set to account for the duration of prolonged PCR positivity of several weeks in these patients. The cutoff determination was further informed by the distribution of the time difference between the first-positive swab and subsequent positive swabs among SARS-CoV-2 cases with multiple swabs (Figure 1). The tail of this distribution indicates that a cutoff of 45 days (at the 99^th^ percentile) provides an appropriate mark for defining the end of prolonged PCR positivity: a subsequent positive swab within 45 days of the first-positive swab is likely to reflect prolonged PCR positivity (due to non-viable virus fragments) rather than reinfection, and thus should not be included in analysis.

**Figure 1.**
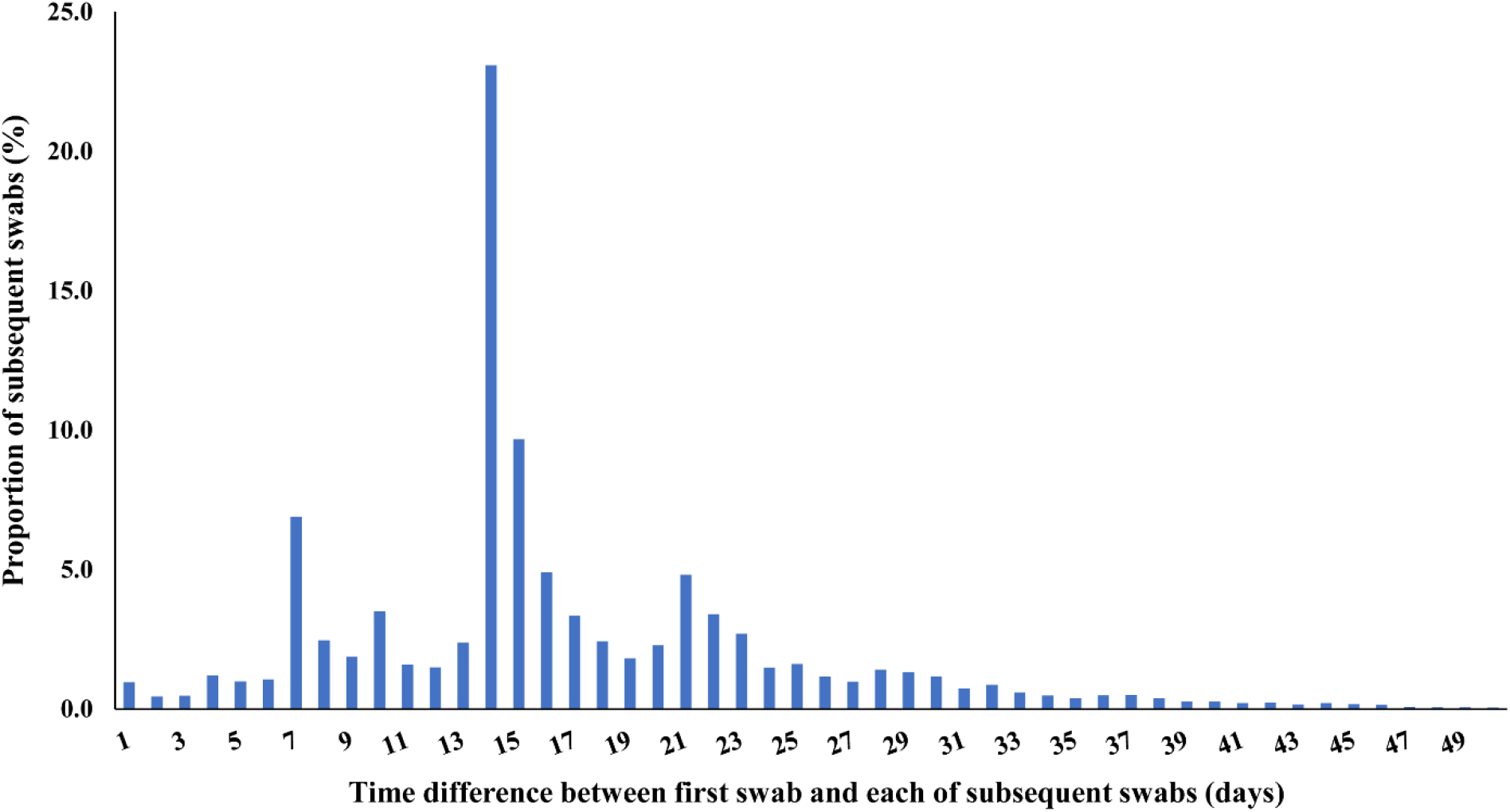
Distribution of the time difference between the first swab and subsequent swabs among all laboratory-confirmed SARS-CoV-2 cases with more than one positive swab. The cutoff of 45 days was at the 99^th^ percentile, and thus provides an appropriate mark for defining the end of the prolonged polymerase chain reaction (PCR) positivity.

### Suspected reinfection case classification

Suspected cases of reinfection, that is cases fitting above indicated inclusion criteria, were classified as showing either *strong* evidence, *good* evidence, *some* evidence, or *weak* (or *no*) evidence for reinfection (Box 1). The classification was based on holistic quantitative and qualitative criteria applied to each investigated case. The criteria included the pattern and magnitude of the change in PCR cycle threshold (Ct) value across repeated swabs, time interval between subsequent swabs, PCR testing site (such as outpatients at primary care, hospital emergency, or inpatient hospitalization), purpose of PCR testing (such as appearance of symptoms, contact tracing, or survey/testing campaign), age, history of COVID-19-related hospital admission, and case severity per WHO classification [12].

Overall, swabs with Ct <30 (suggestive of recent active infection) at least 45 days after the first-positive swab were considered as showing *strong evidence for reinfection*. Swabs with Ct ≥30 at least 45 days after the first-positive swab were considered as showing *good evidence for reinfection* if PCR positivity was associated with *contextual evidence* supporting the status of “reinfection” including appearance of symptoms (often as proxied by being diagnosed at a health facility), if the infection was diagnosed through contact tracing (indicating recent exposure to an infected person), if the change in Ct value from the last swab was to a lower Ct value (indicating increasing viral load), and/or if the repeated swabbing did not follow a regular pattern and time interval between repeated swabs was not short (to exclude cases under clinical management that are indicative of poor control of first infection).

Shorter durations bordering the cutoff of 45 days with Ct values ≥30 and with no contextual evidence supporting the status of “reinfection” were indicative of *some evidence for reinfection*, but not strong nor good evidence for reinfection, as they are more likely to reflect the long tail of the prolonged PCR positivity distribution (Figure 1) [14, 15]. Age ≥70 years, repeated swabs on hospitalized patients, and severe or critical WHO disease classifications were considered as contextual factors indicative of poor infection control of the first infection rather than reinfection. Cases that had such contextual factors (and implicitly did not fit the criteria of strong, good, or some evidence for reinfection) were considered to have *weak (or no) evidence for reinfection*.

Of note that hospitalized COVID-19 cases often had multiple subsequent swabs administered to them as part of clinical care, and repeated swabbing was standard earlier in the epidemic, as the criteria for discharge from an isolation facility required at least two subsequent PCR negative swabs. This was changed later on to a time-based criteria per updated WHO recommendation [16].

### Reinfection risk and rate

Documented reinfection *risk* was assessed by quantifying the proportion of cases with *strong or good evidence for reinfection* out of all laboratory-confirmed SARS-CoV-2 cases. *Incidence rate* of documented reinfection was calculated by dividing the number of cases with strong or good evidence by the number of person-weeks contributed by all laboratory-confirmed cases who had their first-positive swab ≥45 days before day of analysis. The follow-up person-time was calculated starting from 45 days after the first-positive swab and up to the reinfection swab, all-cause death, or end-of-study censoring.

### Sensitivity analyses

Since we implemented a conservative approach prone to overestimate risk of documented reinfection, several sensitivity analyses were conducted implementing more *stringent criteria* for assessing reinfection: 1) exclusion of cases where the Ct value for the first and/or subsequent positive swab was unknown or with a value ≥35 (to exclude potential PCR false-positive cases), 2) changing the ≥45-day cutoff to a ≥60-day cutoff to further exclude potential cases of long-term prolonged PCR positivity, and (*most stringent*) 3) setting definition of recent active infection at Ct cutoff value of <25 (instead of <30) and excluding any suspected reinfection case with Ct value >25.

### Viral genome sequencing and analysis

Viral genome sequencing was conducted on retrieved paired samples of the first-positive swab and reinfection swab for patients with *strong* or *good evidence for reinfection* as confirmatory analysis. Viral RNA was extracted using Quick-RNA Viral Kit (Zymo Research, Irvine, USA; Cat. No. R1041) and eluted in 30ul of nuclease-free water. RNA quality was assessed with RT-qPCR using SARS-CoV-2 (2019-nCoV) CDC qPCR Probe Assay Research Use Only (RUO) kit (Integrated DNA Technologies, USA; Cat number 10006713) and Luna Universal Probe One-Step RT-qPCR Kit (New England BioLabs, USA; Cat number E3006E) on an Applied Biosystems 7500 Fast Real-Time PCR instrument (Applied Biosystems, CA, USA).

Next-generation sequencing (NGS) library construction was performed using the CleanPlex SARS-CoV-2 Panel (Paragon Genomics, USA; SKU: 918012). Gel-size selection on a 3% agarose gel was further utilized to prevent formation of adapter dimers. NGS libraries were then quantified using KAPA Library Quantification Kit (Roche, USA; KK4824), and normalized, pooled, and sequenced on an Illumina MiSeq instrument using a paired-end 150bp kit (Illumina, USA; MS-102-2002). All procedures were implemented following manufacturers’ protocols.

Raw sequences were processed with CUTADAPT (v2.10) [17] to exclude the contaminating adapter sequences. Adapter trimming was performed using the parameters -g CCTACACGACGCTCTTCCGATCT **-a** AGATCGGAAGAGCACACGTCTGAA **-A** AGATCGGAAGAGCGTCGTGTAGG **-G** TTCAGACGTGTGCTCTTCCGATCT **-e** 0.1 **-O** 9 **-m** 50 **-n** 2. Only paired reads with minimum length of 50bp were retained for analysis. The latter filtered reads were aligned to SARS-CoV-2 reference genome (NC_045512) using BWA-MEM [18]. FGBIO (v1.3.0) was subsequently used to remove PCR primer sequences from the resulting BAM file.

Variant calling and genotyping were performed with VarScan multi-sample mpileup [19] with the pileup file generated using SAMTOOLS mpileup (v1.10) [20] with --min-BQ 20 and --min-MQ 20 parameters. The mpileup2snp function of VarScan was then applied with the filtering parameters --min-var-freq 0.2, --min-coverage 5, and --min-avg-qual 20, to generate the final VCF file.

### Ethical approval

The study was approved by HMC and Weill Cornell Medicine-Qatar Institutional Review Boards.

## RESULTS

### Epidemiological analysis

Figure 2 illustrates the selection process of SARS-CoV-2 eligible cases and summarizes the results of their reinfection status’ evaluation. Out of 133,266 laboratory-confirmed cases, 117,458 had only one single positive swab and thus were excluded from further analysis. Of the remaining 15,808 cases with multiple swabs, only 243 persons had *at least* one subsequent positive swab that is ≥45 days from the first-positive swab, and thus qualified for inclusion in analysis.

**Figure 2.**
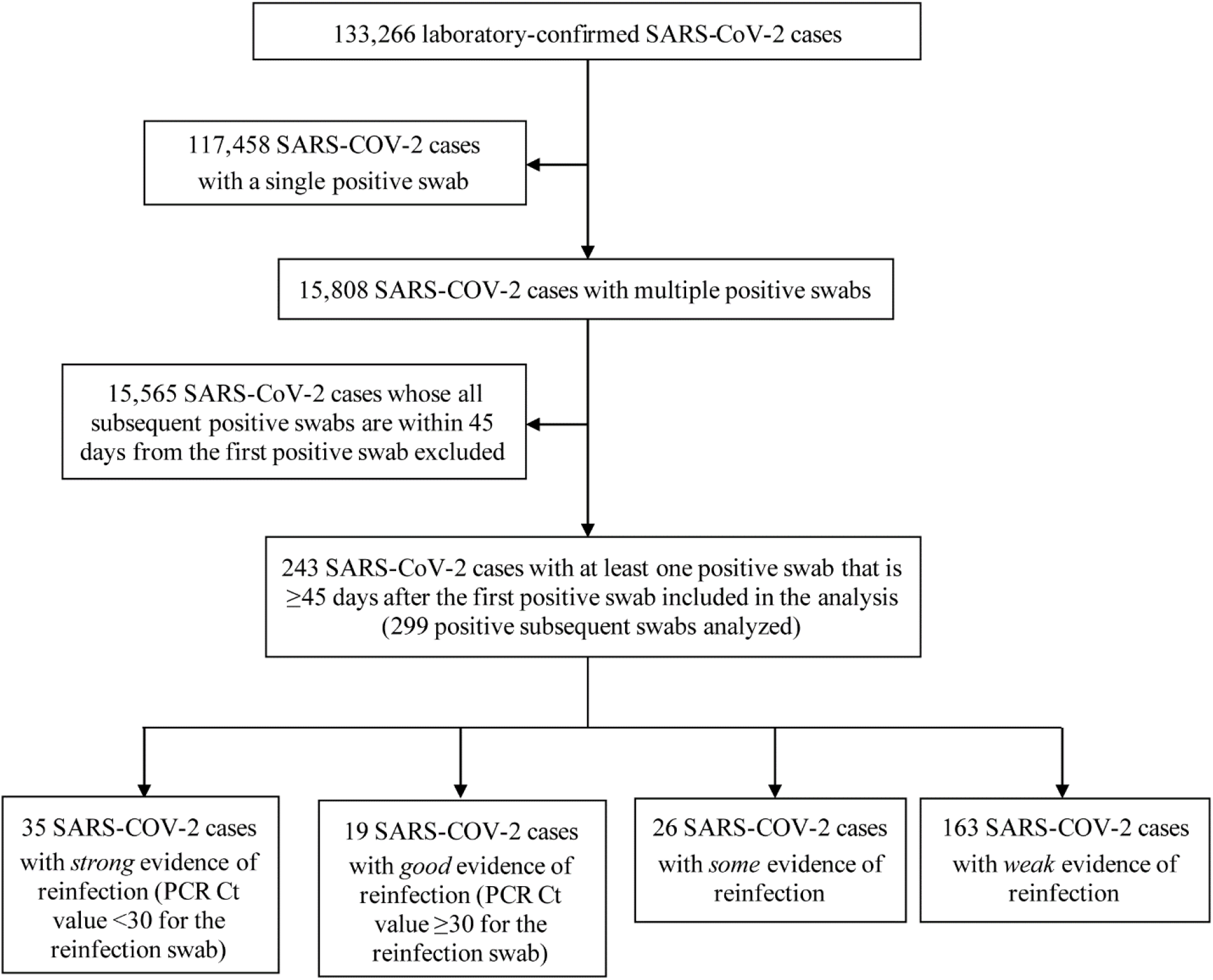
Flow chart describing the selection process of SARS-CoV-2 eligible cases and summarizing the results of their reinfection status’ evaluation.

There were 299 positive swabs collected ≥45 days after the first-positive swab for these 243 persons. Individual investigation of each of these swabs yielded 54 cases with *strong* or *good* evidence for reinfection. Of these, 35 had *strong* evidence for reinfection (Ct value <30) while the remaining 19 had *good* evidence for reinfection (Ct value ≥30). An additional 26 cases showed *some* evidence for reinfection, while evidence was *weak* for the remaining 163 cases.

Table 1 shows the characteristics of the 54 cases classified as showing strong or good evidence for reinfection. Almost all cases were males, but this reflects the focus of the epidemic in craft and manual workers [6]. The median age was 33 years (range: 16-57) and the median time between the *first* swab and the *reinfection* swab was 64.5 days (range: 45-129). The median Ct value was 28 (range: 14-37): it was 22 (range 14-29) for the 35 swabs classified with strong evidence (Ct <30) and 32 (range: 30-37) for the remaining swabs (Ct ≥30).

**Table 1.**
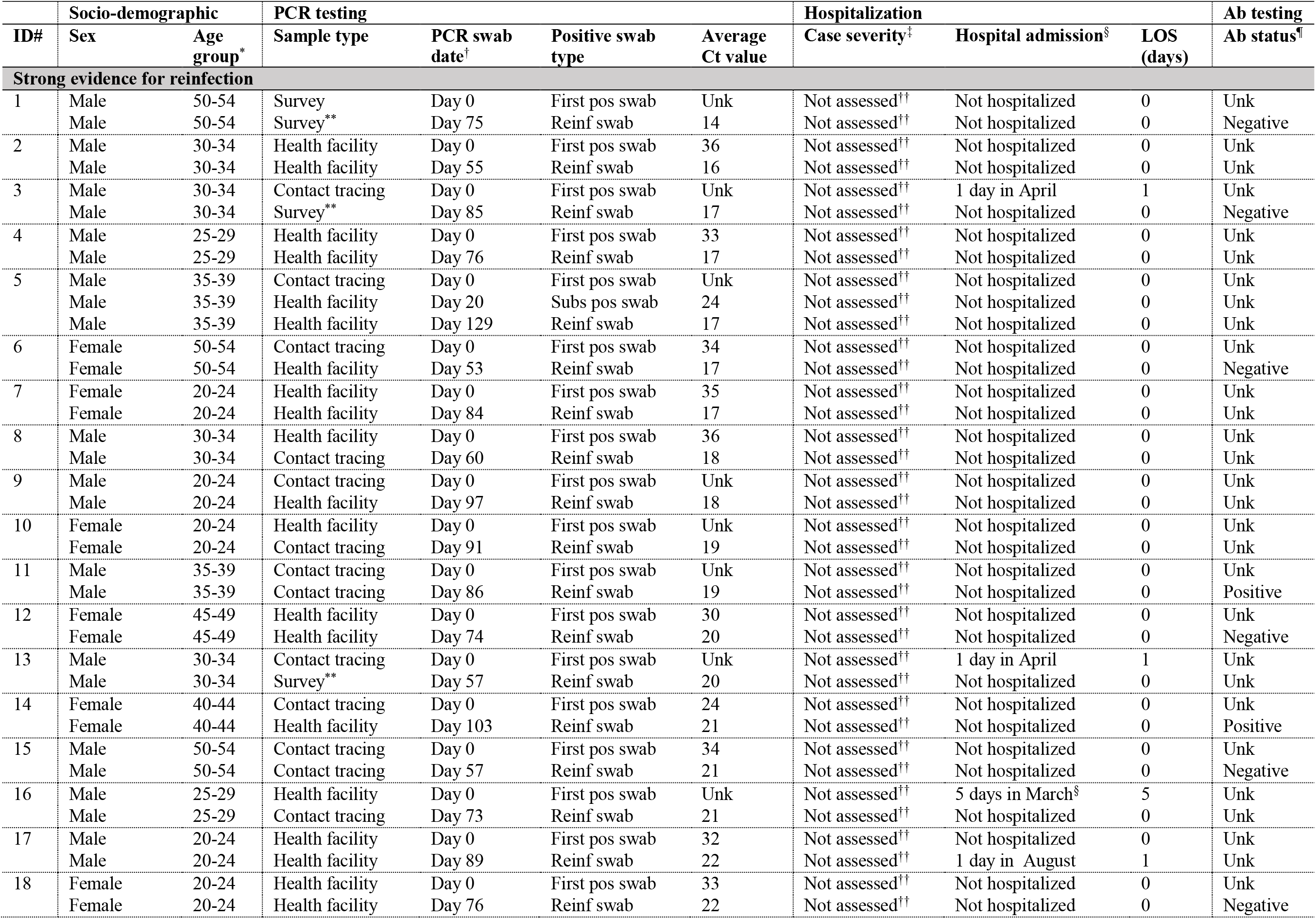
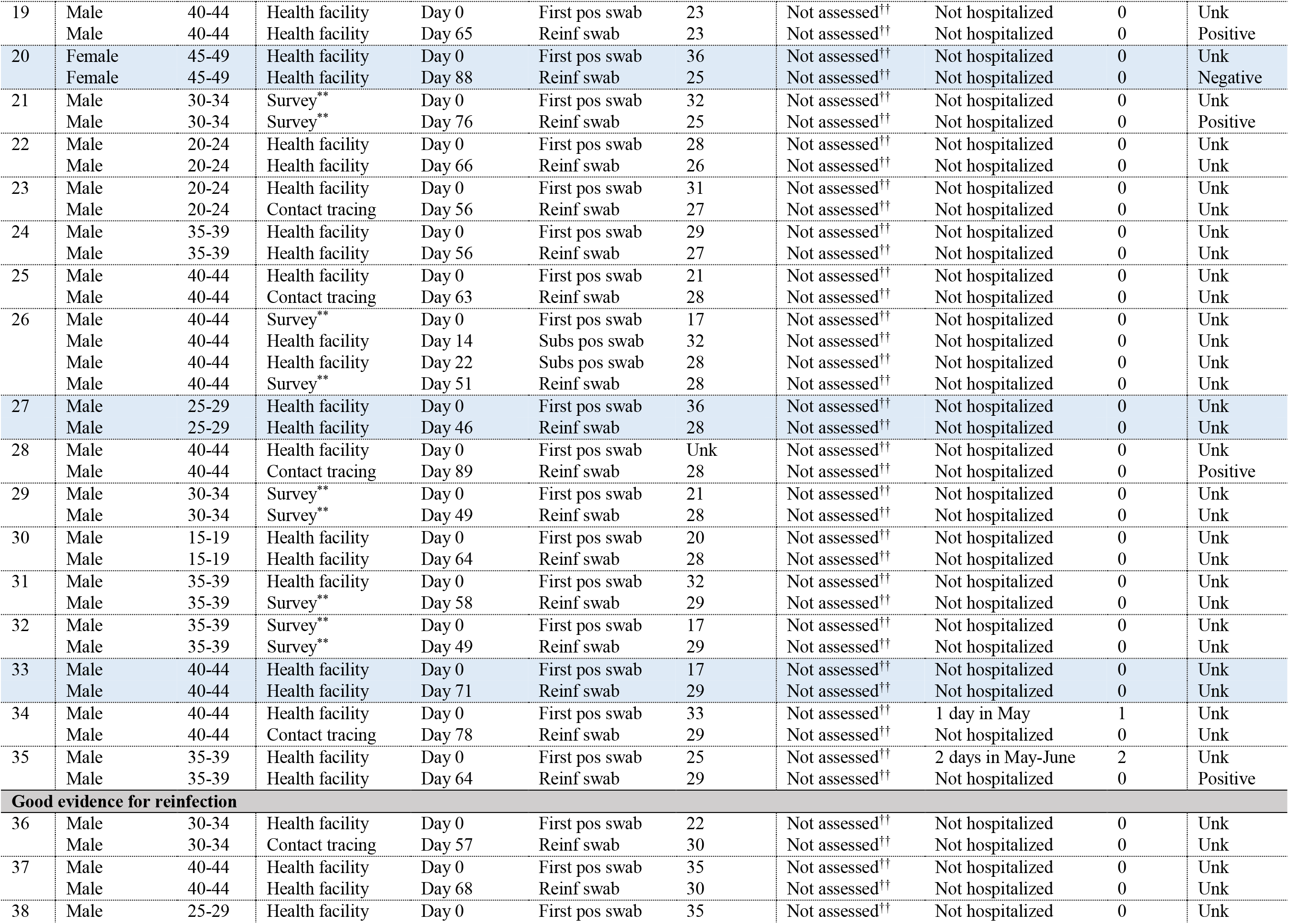
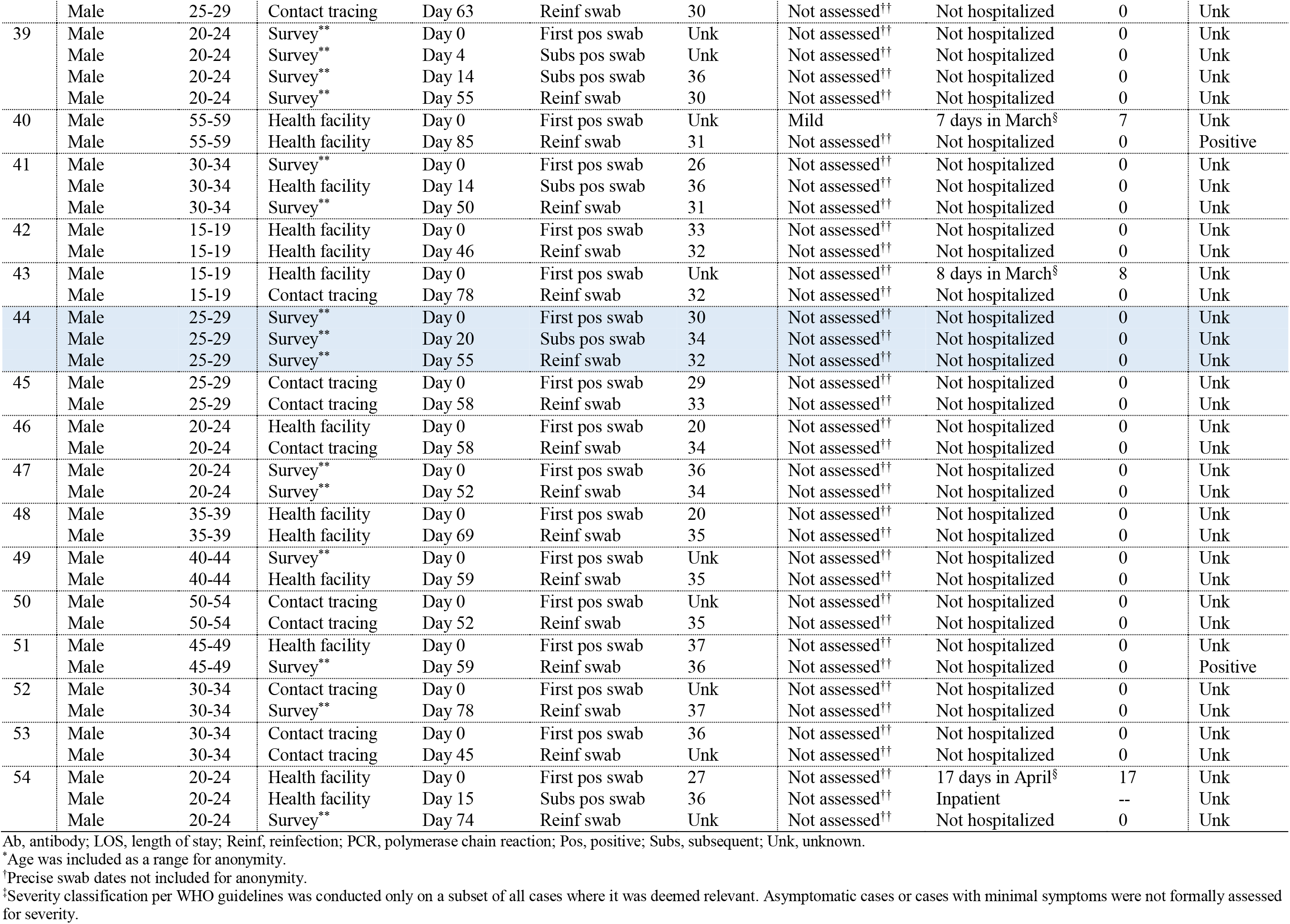
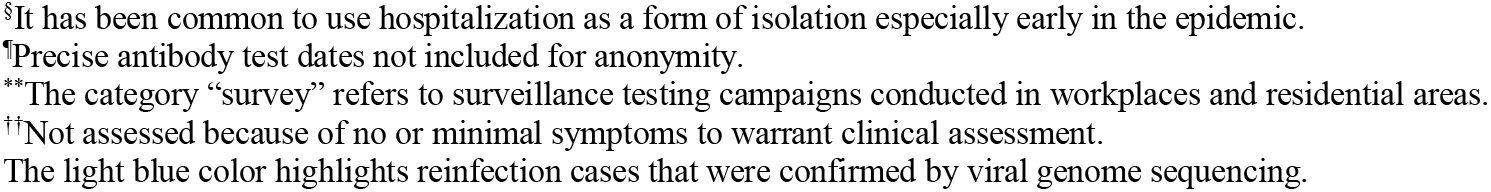
Characteristics of individuals classified as showing strong or good evidence for reinfection.

Twenty-three cases (42.6%) were diagnosed at a health facility, suggesting presence of symptoms while 31 (57.4%) were identified incidentally either through random testing campaigns/surveys (n=15; 27.8%) or contact tracing (n=16; 29.6%), suggesting minimal symptoms if any.

Nine cases were hospitalized at any time, all but *one* occurred following the first infection episode and mostly for isolation purposes. Only one case had sufficient symptoms to warrant clinical assessment (during primary infection), but was classified with “mild” severity per WHO classification. No deaths were recorded. Of note that the vast majority of infections in Qatar occurred in young and healthy men and had limited severity [6].

Antibody test results were available for 48 out of the 243 assessed individuals (Table 2), of whom 30 (62.5%) had detectable antibodies. Of the 13 with strong evidence for reinfection *and* available antibody results, seven (53.9%) were sero-negative. Meanwhile, both individuals with good evidence for reinfection, three of the four individuals with some evidence for reinfection, and 19 of the 29 individuals with weak evidence for reinfection, were sero-positive.

**Table 2.** Distribution of cases according to reinfection and antibody statuses (for individuals with complete information for these statuses).

| **Reinfection status** | **Seronegative** | **Seropositive** | **Total** |
| --- | --- | --- | --- |
|  | N (%) | N (%) | N (%) |
| **Strong evidence for reinfection** | 7 (53.9) | 6 (46.2) | 13 (100.0) |
| **Good evidence for reinfection** | 0 (0.0) | 2 (100.0) | 2 (100.0) |
| **Some evidence for reinfection** | 1 (25.0) | 3 (75.0) | 4 (100.0) |
| **Weak evidence for reinfection** | 10 (34.5) | 19 (65.5) | 29 (100.0) |
| **Total N (%)** | 18 (37.5) | 30 (62.5) | 48 (100.0) |

Risk of documented reinfection was estimated at 0.04% (95% CI: 0.03-0.05%)—that is a total of 54 reinfections in the cohort of 133,266 laboratory-confirmed SARS-CoV-2 infected persons. Incidence rate of reinfection was estimated at 1.09 (95% CI: 0.84-1.42) per 10,000 person-weeks—that is a total of 54 reinfection events in a follow-up person-time of 495,208.7 person-weeks.

The results of the sensitivity analyses can be found in Appendix Table S1. In these analyses, the estimate for the risk of reinfection ranged between 0.01% (95% CI: 0.01-0.02) and 0.02% (95% CI: 0.02-0.03), while the estimate for the incidence rate of reinfection ranged between 0.38 (95% CI: 0.24-0.60) and 1.06 (95% CI: 0.75-1.50) per 10,000 person-weeks. Although these sensitivity analyses confirmed our results, they suggested that we may have overestimated the already low risk of reinfection.

### Confirmation of reinfection through viral genome sequencing

Paired specimens of the first-positive and reinfection swabs could be retrieved for 23 out of the 54 cases with strong or good evidence for reinfection. Table 3 summarizes the viral genome sequencing results and Figure 3 and Appendix Figures S1-S2 show the detailed analysis for each genome pair.

**Table 3.** Results of reinfection confirmatory analysis based on viral genome sequencing of the paired viral specimens of the first-positive and reinfection swabs for 23 patients with strong or good epidemiological evidence for reinfection.

| **Viral genome sequencing evidence for reinfection** | **Indication upon comparing each genome pair** | **N** |
| --- | --- | --- |
| Insufficient evidence to warrant interpretation | One or two genomes of low quality | 11 |
| No evidence for reinfection | One change of allele frequency | 3 |
| Shifting balance of quasi-species with no evidence for reinfection | Several changes of allele frequency | 3 |
| Conclusive evidence for no reinfection | Both genomes of high quality yet no differences found | 2 |
| Supporting evidence for reinfection | One genome of inferior quality but with D614G mutation | 2 |
| Conclusive evidence for reinfection | Multiple changes of allele frequency and D614G mutation | 2 |
| **Total** |  | **23** |

**Figure 3.**
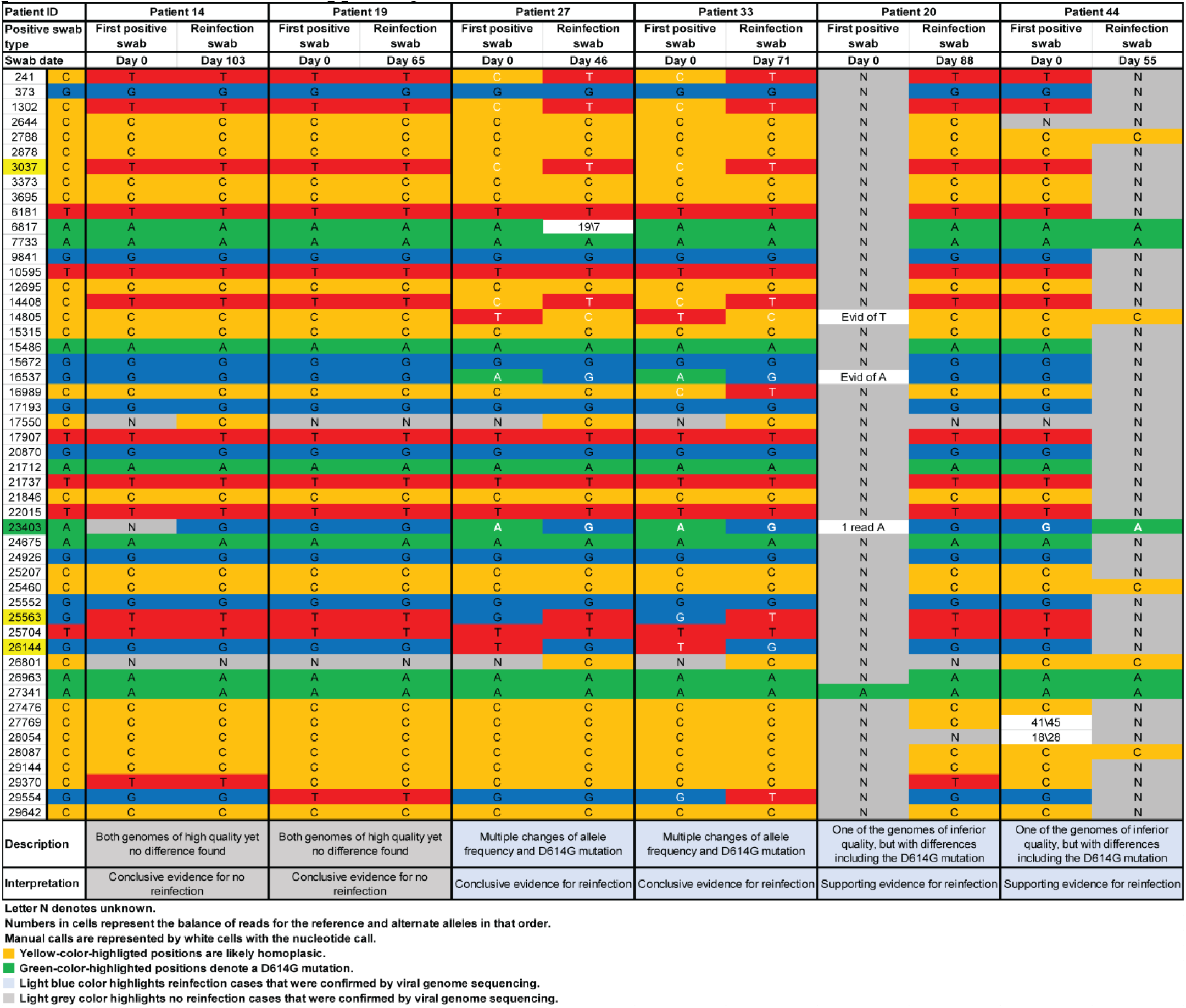
Viral genome sequencing analysis of the paired viral specimens of the first-positive and reinfection swabs for the six patients with conclusive or supporting evidence for reinfection or no reinfection.

There was insufficient evidence to warrant interpretation for 11 pairs because of low genome quality. For six pairs, there were one to several changes of allele frequency indicative at best of a shifting balance of quasi-species, and thus no evidence for reinfection. For two pairs, remarkably, there was conclusive evidence for *no reinfection* as both genomes were of high quality yet no differences were found. For both patients, the Ct value was <25 for the first-positive and reinfection swabs indicating persistent active infection (Table 1). These two cases were also sero-positive (Table 1).

Meanwhile, for two pairs, there was conclusive evidence for reinfection with multiple changes of allele frequency and presence of the D614G mutation (23403bp A>G)—a variant that appeared and expanded replacing the original D614 form [21, 22]. Also for two pairs, and although one of the genomes was of inferior quality, there was sufficient evidence for differences including the presence of the D614G mutation, thereby rendering evidence for reinfection. Three out of these four cases with viral genome sequencing confirmation of reinfection were classified above (epidemiological criteria) as having strong evidence for reinfection, with the fourth classified as having good evidence (Table 1). Antibody test result was available for one case at time of reinfection, and the individual was sero-negative.

In sum, for the 12 cases where viral genome sequencing evidence was available, four cases were confirmed as reinfections, a confirmation rate of 33.3%. Applying this rate to the above-estimated reinfection metrics yielded risk of documented reinfection of 0.01% (95% CI: 0.01-0.02%) and incidence rate of reinfection of 0.36 (95% CI: 0.28-0.47) per 10,000 person-weeks.

## DISCUSSION

Results indicate, employing several analyses and sensitivity analyses, conclusive evidence for presence of reinfections in the SARS-CoV-2 epidemic of Qatar, but the risk for documented reinfection was very rare at about 1-2 reinfections per 10,000 infected persons. This finding is striking as the epidemic in Qatar has been intense with half of the population estimated to have been infected [6]. Considering the strength of the force of infection, estimated at a *daily* probability of infection exceeding 1% at the epidemic peak around May 20 [6], it is all but certain that a significant proportion of the population has been repeatedly exposed to the infection, but such re-exposures hardly led to any documentable reinfections.

Indeed, of all epidemiologically-identified reinfections, nearly two-thirds (57%) were discovered accidentally, either through random testing campaigns/surveys or through contact tracing. None were severe, critical, or fatal, all reinfections were asymptomatic or with minimal or mild symptoms. These findings suggest that most infected persons do develop immunity against reinfection that lasts for at least few months, and that reinfections (if they occur) are well tolerated and no more symptomatic than primary infections. Further follow up of this cohort of infected persons over time may allow elucidation of any potential effects of waning of immunity.

Other lines of evidence for this cohort also support this conclusion. Among 2,559 PCR positive persons where an antibody test outcome was available [6], and where the first-positive PCR test was conducted >3 weeks before the serology test to accommodate for the delay in development of antibodies following onset of infection [14, 15], 91.7% were antibody positive [6]. The high antibody positivity was also stable for over three months [6], as described elsewhere [9]. The epidemic curve in Qatar was further characterized by rapid growth followed by rapid decline [6], at a time when levels of social and physical distancing restrictions were fairly stable. This points to susceptibles-infected-recovered “SIR” epidemic dynamics with most infections eliciting immunity against reinfection.

This assessment has limitations. We assessed risk of only *documented* reinfections, but other reinfections could have occurred but went undocumented, perhaps because of minimal or no symptoms. It is possible that with the primed immune system following the primary infection, reinfections are milder and shorter and thus less likely to be documented [10], though still can be infectious as the Ct value was quite low, indicating high viral load, in some of the reinfections. Viral genome sequencing analysis was possible for only a subset of reinfections. Antibody testing outcomes were available for only a number of cases, limiting use and inferences of the link between antibody status and risk of reinfection. Of note that for one of the genetically-confirmed reinfections the antibody test result was available but was sero-negative, just as the Hong Kong reinfected patient [23].

In conclusion, SARS-CoV-2 reinfection appears to be a rare phenomenon. This suggests that immunity develops after the primary infection and lasts for at least a few months, and that immunity protects against reinfection.

#### Box 1. Classification of suspected cases of SARS-CoV-2 reinfection based on strength of supporting epidemiological evidence.

Suspected cases of SARS-CoV-2 reinfection: all laboratory-confirmed cases with at least one polymerase chain reaction (PCR) positive swab that is ≥45 days after a first-positive swab.

*Strong* evidence for reinfection: individuals having positive swabs with PCR cycle threshold (Ct) value <30 at least 45 days after the first-positive swab. No contextual evidence supporting poor control of first infection such as age ≥70 years, repeated swabs on hospitalized patients, and severe or critical World Health Organization disease classifications.

*Good* evidence for reinfection: individuals having positive swabs with PCR Ct value ≥30 at least 45 days after the first-positive swab, but where PCR positivity was associated with contextual evidence supporting the status of reinfection:

- Appearance of symptoms (often as proxied by being diagnosed at a health facility)
- Infection diagnosis through contact tracing (indicating recent exposure to an infected person)
- Lower Ct value compared to last positive swab (indicating increasing viral load)
- Irregular and spaced-out pattern for repeated swabbing (to exclude cases under clinical management that are indicative of poor control of first infection).

No contextual evidence supporting poor control of first infection such as age ≥70 years, repeated swabs on hospitalized patients, and severe or critical World Health Organization disease classifications.

*Some* evidence for reinfection: individuals having positive swabs with PCR Ct value ≥30 at least 45 days after the first-positive swab, but typically bordering the cutoff of 45 days. PCR positivity was **not** associated with evidence supporting the status of reinfection (listed above).

*Weak* evidence for reinfection: individuals having swabs with PCR Ct value ≥30 at least 45 days after the first-positive swab, but typically bordering the cutoff of 45 days. PCR positivity was associated with contextual evidence indicative of poor infection control of the first infection rather than reinfection (such as age ≥70 years, repeated swabs on hospitalized patients, and severe or critical World Health Organization disease classifications).

## Data Availability

Relevant data are available within the manuscript and appendix.

## Acknowledgements

We would like to thank Her Excellency Dr. Hanan Al Kuwari, the Minister of Public Health, for her vision, guidance, leadership, and support. We also would like to thank Dr. Saad Al Kaabi, Chair of the System Wide Incident Command and Control (SWICC) Committee for the COVID-19 national healthcare response, for his leadership, analytical insights, and for his instrumental role in enacting the data information systems that made these studies possible. We further extend our appreciation to the SWICC Committee and the Scientific Reference and Research Taskforce (SRRT) members for their informative input, scientific technical advice, and enriching discussions. We would also like to thank Dr. Mariam Abdulmalik, the CEO of the Primary Health Care Corporation and the Chairperson of the Tactical Community Command Group on COVID-19, as well as members of this committee, for providing support to the teams that worked on the field surveillance. We also would like to acknowledge the dedicated efforts of the Clinical Coding Team and the COVID-19 Mortality Review Team, both at Hamad Medical Corporation, and the Surveillance Team at the Ministry of Public Health.

## Author contributions

LJA conceived and co-designed the study and led the statistical analyses. HC co-designed the study, performed the data analyses, and wrote the first draft of the article. JAM led the viral genome sequencing analyses and AAA, YAM, and SY conducted these analyses. All authors contributed to data collection and acquisition, database development, discussion and interpretation of the results, and to the writing of the manuscript. All authors have read and approved the final manuscript.

## Appendix

**Table S1.** Sensitivity analyses assessing the robustness of our estimates for the risk of reinfection and the incidence rate of reinfection to more stringent criteria for determining reinfection.

| Analysis type | Risk of reinfection | Incidence rate of reinfection |
| --- | --- | --- |
|  | Estimate (95% CI) in % | Estimate (95% CI) per 10,000 person-weeks |
| **Sensitivity analysis** |  |  |
| Excluding all cases where PCR cycle threshold (Ct) value for the first and/or subsequent positive swab was unknown or with a value $\geq 35$ (to exclude potential PCR false-positive cases) | 0.02 (95% CI: 0.01-0.03) | 0.52 (95% CI: 0.36-0.77) |
| Changing the $\geq 45$-day cutoff to a $\geq 60$-day cutoff to further exclude potential cases of long-term prolonged PCR positivity | 0.02 (95% CI: 0.02-0.03) | 1.06 (95% CI: 0.75-1.50) |
| Excluding any suspected reinfection with Ct value $> 25$ | 0.01 (95% CI: 0.01-0.02) | 0.38 (95% CI: 0.24-0.60) |
| **Main analysis** | 0.04 (95% CI: 0.03-0.05) | 1.09 (95% CI: 0.84-1.42) |

CI, confidence interval; PCR, polymerase chain reaction.

**Figure S1.**
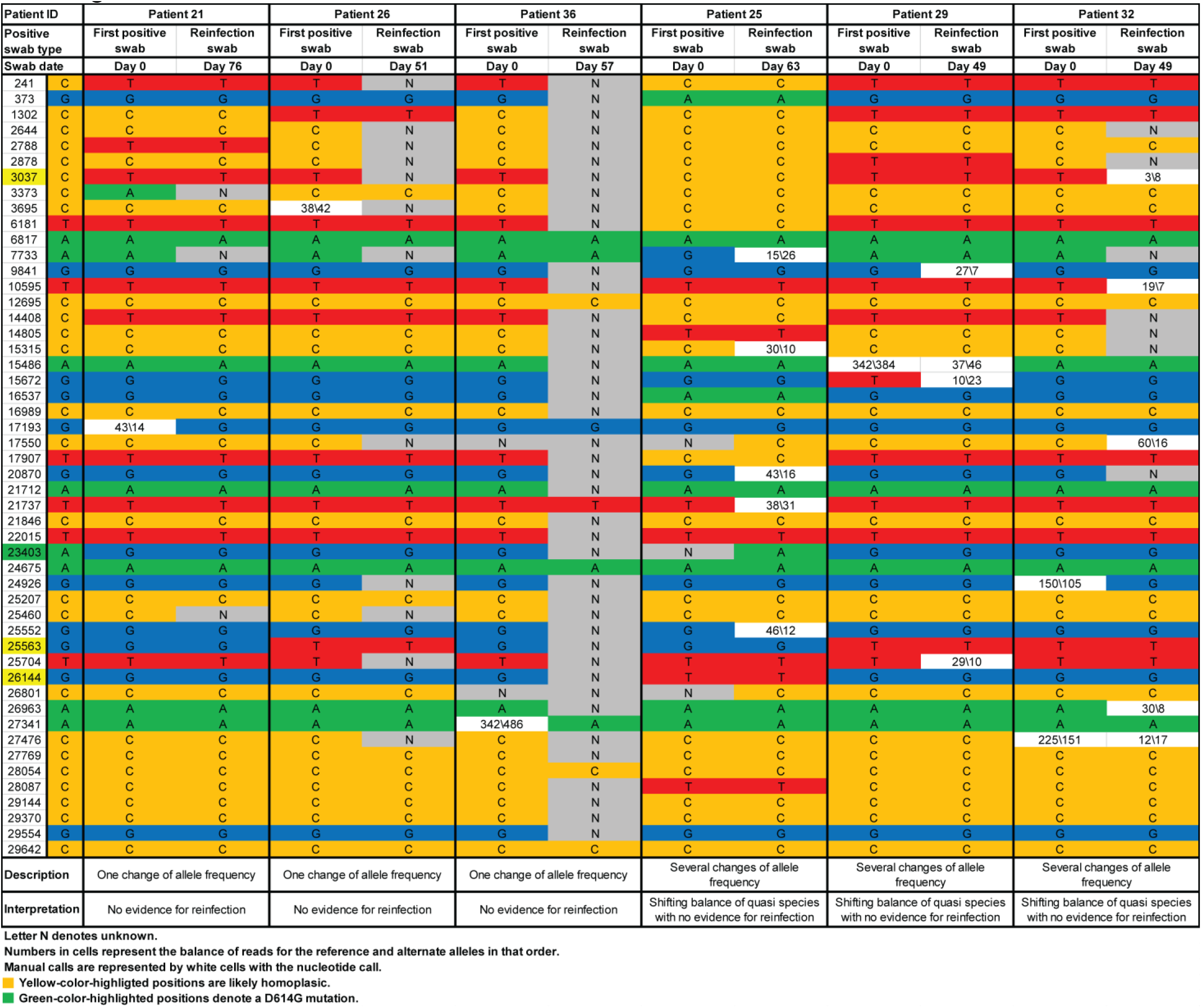
Genetic sequencing analysis of the paired viral specimens of the first-positive and reinfection swabs for the six patients with no genetic evidence for reinfection.

**Figure S2.**
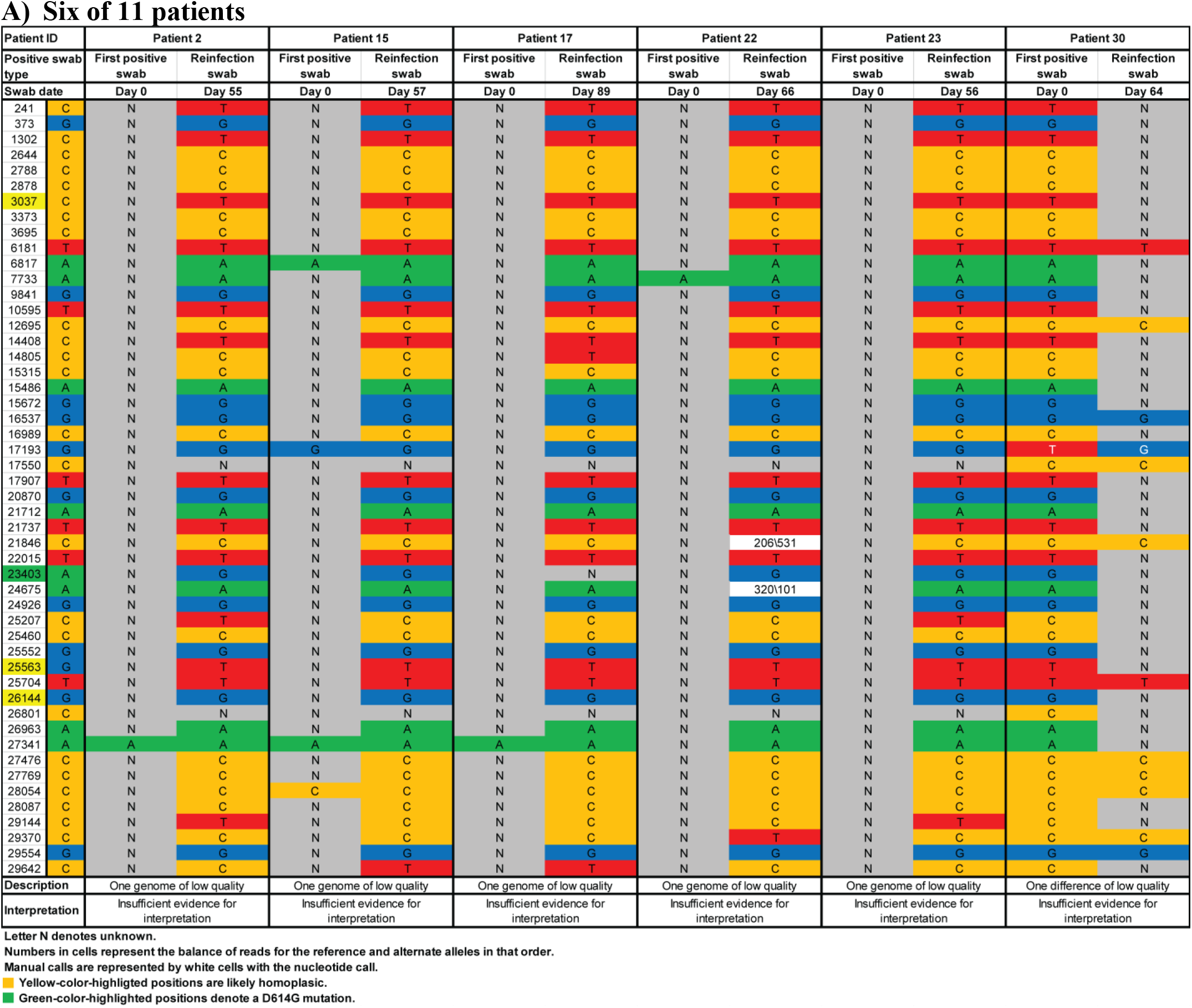
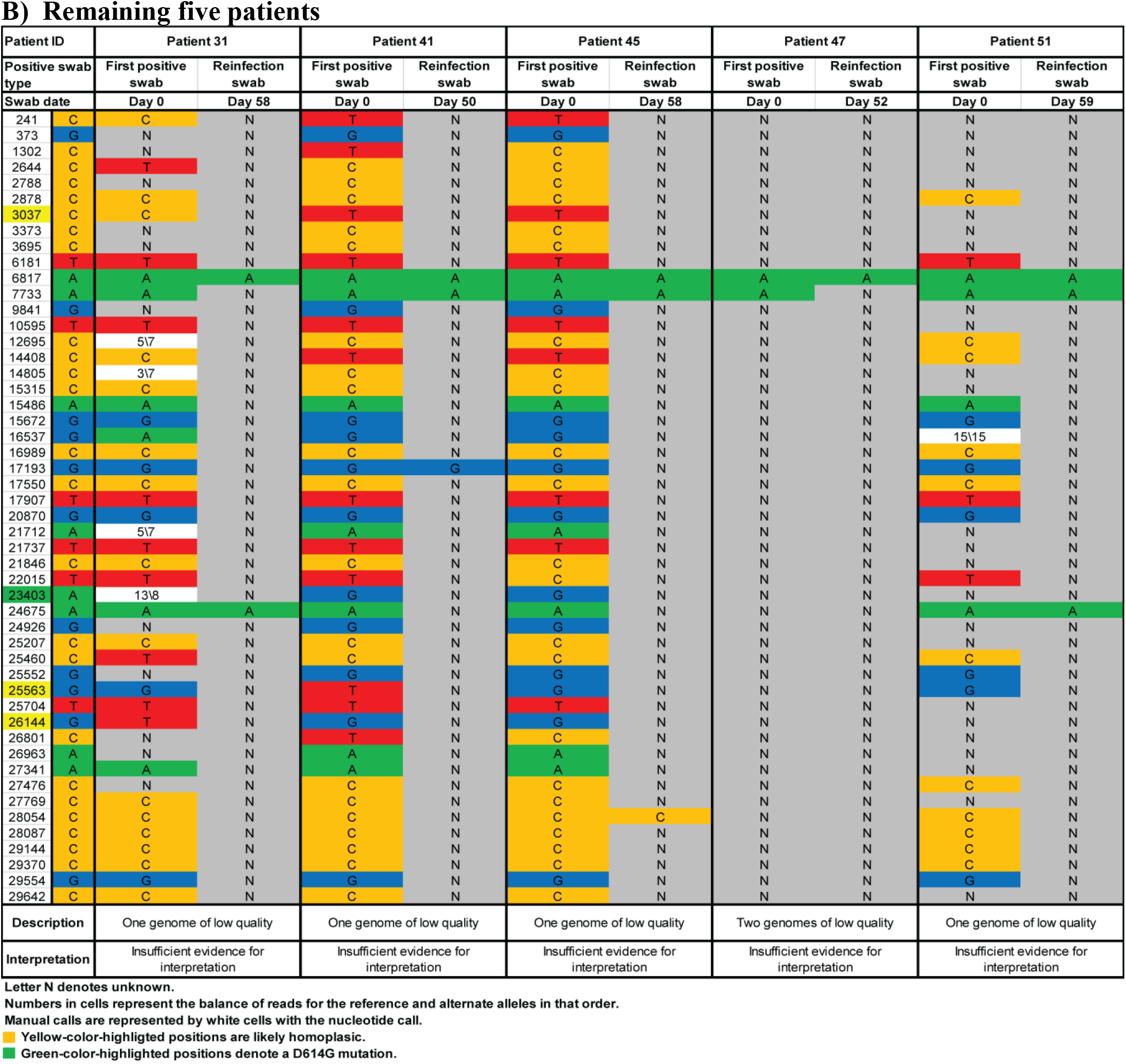
Genetic sequencing analysis of the paired viral specimens of the first-positive and reinfection swabs for the cases with insufficient genetic evidence to confirm reinfection.

